# Comparing epigenetic puberty and epigenetic clocks as biomarkers of adult biological aging and health

**DOI:** 10.64898/2026.09.29.26364286

**Authors:** Q. Wu, S.F.P. Gibbs, A.J. de Steiguer, S. Zarandooz, Y.E. Willems, T. Smith, J.A. Goode, C. Mitchell, E. M. Crimmins, K. P. Harden, R.E. Marioni, L. Raffington

**Author notes:** **Corresponding Author:** Dr. Laurel Raffington, Max Planck Institute for Human Development; Max Planck Research Group Biosocial – Biology, Social Disparities, and Development; Lentzeallee 94, 14195 Berlin, Germany.

## Abstract

Recent lifespan frameworks of aging theorize that pubertal development and cellular aging are governed by overlapping pathways, but the time lag between adolescence and older adulthood complicates the study of shared molecular mechanisms. Here we compare a DNA-methylation-based epigenetic index of pubertal development to epigenetic clocks, which were designed to measure biological aging, as a putative biomarker of adult aging and health in two adult cohorts: 3,642 older Americans in the Health and Retirement Study (HRS; age range, 56 to 100 years; 2,106 females) and 18,676 adults from Generation Scotland (GS; age range, 18 to 98 years; 10,974 females). Epigenetic puberty was correlated with chronological age in both cohorts, moderately in the older HRS sample (r = 0.51 in females, r = 0.51 in males) and strongly across the wider adult age range of GS (r = 0.82 in females, r = 0.86 in males). In both cohorts, age-residualized epigenetic puberty was associated with accelerated biological aging as measured by epigenetic clocks and other aging biomarkers (standardized coefficients: HRS, 0.13 to 0.47, all P < 0.005; GS, 0.05 to 0.40, all P < 0.001). Epigenetic puberty was linked with multiple health outcomes: mortality in HRS in both sexes (ORs, 1.39 and 1.46, both P ≤ 0.001) and in GS among females (HR, 1.12, P = 0.003); 10-year onset of diabetes, any cancer and lung disease in GS (HRs, 1.12 to 1.19, all P < 0.005); and with multimorbidity, functional limitations and BMI in females in HRS (standardized coefficients, 0.10 to 0.16, all P < 0.001). It also accounted for a unique, albeit smaller, proportion of variance in aging-related outcomes beyond that explained by epigenetic clocks. These findings indicate that the epigenetic puberty index reflects pro-aging underlying biology, showing overlapping molecular pathways between pubertal development and cellular aging, and support a lifespan-based approach to studying biological aging.

## Introduction

Recent frameworks posit that cellular biological aging is a lifelong process beginning well before adulthood (Gladyshev, 2021; Horvath & Raj, 2018; Raffington, 2024). Accordingly, there is growing interest in generating and tracking biomarkers of development and biological aging from infancy onward (e.g. PROSPER Consortium, Strauss et al., 2026). Pubertal development is one such early-life window that may share molecular mechanisms with later-life aging (Horvath & Raj, 2018; Lu et al., 2023; Raffington, 2024). This multi-system transition requires a high allocation of energetic resources toward growth, hormone synthesis, and reproductive maturation that may come at the expense of somatic maintenance and repair (Belsky & Shalev, 2016; Ellis & Giudice, 2019). Moreover, under harsh environmental conditions, accelerated pubertal development is theorized to maximize immediate reproductive success while ultimately compromising long-term health and lifespan (Belsky & Shalev, 2016; Ellis & Giudice, 2019).

Empirically testing potentially shared molecular pathways in humans remains challenging, due to the decades-long time lag between adolescence and older adulthood. Evidence that pubertal development is linked to aging-related health most commonly derives from retrospective studies of middle-aged and older adults. For instance, in adult females, the recall of a younger age at menarche is associated with cardiovascular diseases, type 2 diabetes, and earlier mortality (Elks et al., 2013; Lakshman et al., 2009). In males, early pubertal timing, proxied by adolescent body mass index (BMI), has been linked to increased risk of type 2 diabetes later in life (Ohlsson et al., 2020). Here, we approach this question using a novel and complementary approach. We measure the molecular changes involved in pubertal development in an older population and test whether pubertal biomarkers capture variation in adult biological aging and health (**Figure 1**).

**Figure 1.**
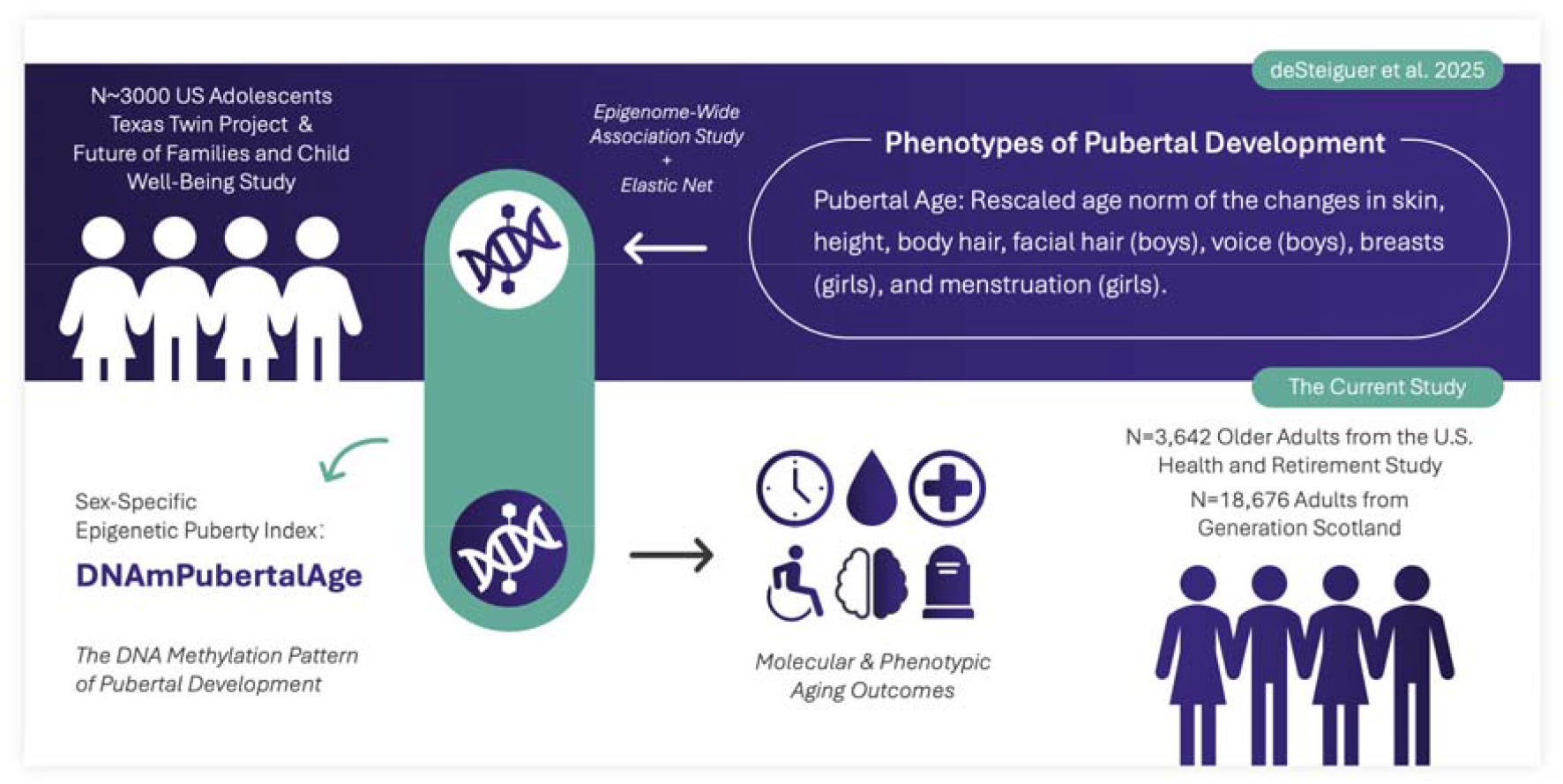
Graphical abstract of the study design.

We build on research on DNA methylation (DNAm) algorithms of biological aging, known as “epigenetic clocks”, which estimate how closely an individual’s DNA methylation patterns resemble those of adults with accelerated physiological aging and elevated mortality risk, relative to healthier, longer lived peers (Raffington & Belsky, 2022). In adults, accelerated epigenetic clocks are robust predictors of aging-related health and mortality (e.g., Faul et al., 2023; Mavrommatis et al., 2025). In adolescents, accelerated epigenetic clocks are associated with earlier pubertal timing (e.g., age at first menses) (e.g., Binder et al., 2018; Raffington et al., 2021; Suarez et al., 2018). However, adolescent samples cannot directly examine links between pubertal development and a full spectrum of aging-related health outcomes, because most such outcomes remain largely unobservable until later adulthood.

Following preregistered analyses (https://doi.org/10.17605/OSF.IO/FXSV7), we applied an epigenetic index of pubertal development, originally developed in over 3000 US adolescents, to two adult cohorts. This index, ‘DNAmPubertalAge,’ was trained to estimate the age at which an individual’s secondary sex characteristics (e.g., breast development, facial hair) align with the population norm (deSteiguer et al., 2025). We calculated the index for n=3642 older adults from the Health and Retirement Study (HRS) and conducted additional unpreregistered analyses in n=18,676 adults from the Generation Scotland (GS) cohort.

Similar to the concept of epigenetic clock acceleration, we quantified ‘DNAmPubertalAge acceleration’ by calculating the residuals from regressing the epigenetic puberty index on chronological age. To determine if the biology captured by this index is relevant to aging, we tested whether it performs similarly to an aging biomarker: a positive correlation with chronological age and associations with multiple interrelated aging outcomes, including epigenetic clocks, physiological dysregulation, multimorbidity, the new onset of aging-related diseases, functioning loss, and mortality. Furthermore, we examined whether epigenetic puberty provides incremental predictive value for downstream aging outcomes after accounting for standard epigenetic clocks.

## Results

### 1. Epigenetic puberty index is associated with aging outcomes in adulthood

We analysed two independent cohorts. The HRS is a nationally representative longitudinal study of middle-aged and older adults aged 51 and above in the United States, and our analytical sample comprised 3,642 respondents (2,106 females and 1,536 males; mean age 68.6 years; age range 56 to 100 years). GS is a population- and family-based Scottish cohort covering a far broader age range (n=18,676; 10,974 females and 7,702 males; mean age 47 years; age range 18 to 98 years). In both cohorts, we computed DNAmPubertalAge, an epigenetic index of pubertal development trained in adolescents, from whole-blood DNA methylation, alongside three established epigenetic clocks (PhenoAge, PCGrimAge and DunedinPACE). Because the epigenetic puberty indices were trained separately for females and males, all the analyses were sex-stratified. The PC version of PhenoAge was not used because HRS was a part of the training sample of PCPhenoAge (Higgins-Chen et al., 2022). Sample characteristics are reported in **Table 1**.

**Table 1.** Sample Characteristics. Mean (standard deviation). HRS: Health and Retirement Study. GS: Generation Scotland. PhenoAge, GrimAge, and DunedinPACE are DNA-methylation-based biological aging measures. AA: Age Acceleration. The AA version of an index was calculated using the residual that resulted from regressing the index on chronological age. PC: Principal Component Version. ExpBioAge: Expanded Biological Age. ExpBioAge is based on clinical biomarkers to measure physiological dysregulation. ADL+IADL: Limitations of Activities of Daily Living and Instrumental Activities of Daily Living. CVD: Cardiovascular Diseases.

|  |  |  |
| --- | --- | --- |
|  | HRS | GS |

**Table 1.** Sample Characteristics.
|  | Females | n | Males | n | Females | n | Males | n |
| --- | --- | --- | --- | --- | --- | --- | --- | --- |
| Age | 69.1 (9.5) | 2106 | 68.0 (8.6) | 1536 | 47.3 (14.8) | 10974 | 46.8 (15.1) | 7702 |
| Epigenetic puberty index |  |  |  |  |  |  |  |  |
| DNAmPubertalAge | 21.2 (2.9) | 2106 | 21.9 (2.1) | 1536 | 17.4 (2.8) | 10974 | 17.4 (2.3) | 7702 |
| DNAmPubertalAgeAA | -0.4 (2.5) | 2106 | 0.5 (1.9) | 1536 | 0.0 (1.6) | 10974 | 0.0 (1.2) | 7702 |
| Epigenetic clocks |  |  |  |  |  |  |  |  |
| PCGrimAge | 76.3 (8.2) | 2106 | 78.5 (7.7) | 1536 | 61.0 (11.4) | 10974 | 62.5 (12.3) | 7702 |
| PCGrimAgeAA | -1.4 (3.5) | 2106 | 1.7 (3.8) | 1536 | -0.8 (3.0) | 10974 | 1.1 (3.2) | 7702 |
| PhenoAge | 56.7 (10.2) | 2106 | 56.9 (9.3) | 1536 | 42.8 (12.0) | 10974 | 42.6 (13.2) | 7702 |
| PhenoAgeAA | -0.5 (6.9) | 2106 | 0.6 (6.7) | 1536 | -0.1 (4.8) | 10974 | 0.1 (4.8) | 7702 |
| DunedinPACE | 1.01 (0.15) | 2106 | 1.04 (0.14) | 1536 | 0.98 (0.13) | 10974 | 0.99 (0.13) | 7702 |
| Aging outcomes in HRS |  |  |  |  |  |  |  |  |
| ExpBioAge | 68.3 (11.5) | 1569 | 68.1 (11.9) | 1139 | - | - | - | - |
| ExpBioAgeAA | -0.2 (7.5) | 1569 | 0.3 (8.7) | 1139 | - | - | - | - |
| Multimorbidity | 0.8 (1.0) | 2106 | 0.9 (0.9) | 1536 | - | - | - | - |
| ADL+IADL | 0.7 (1.6) | 2106 | 0.5 (1.4) | 1536 | - | - | - | - |
| Cognitive Dysfunction | 11.3 (4.5) | 2106 | 11.7 (4.1) | 1536 | - | - | - | - |
| Cognitive Decline | 0.3 (4.0) | 1688 | 0.4 (3.9) | 1166 | - | - | - | - |
| Number of 6-yr mortality cases | 363 | 2091 | 328 | 1525 | - | - | - | - |
| BMI | 29.4 (7.1) | 2089 | 29.4 (6.0) | 1530 | - | - | - | - |
| Earlier menopause | 0.43 (5.24) | 1156 | - | - | - | - | - | - |
| 10-year incident cases in GS |  |  |  |  |  |  |  |  |
| Diabetes | - | - | - | - | 255 | 10818 | 268 | 7513 |
| Cancers (any) | - | - | - | - | 596 | 10575 | 439 | 7524 |
| Lung disease | - | - | - | - | 265 | 10836 | 200 | 7622 |
| Any CVD | - | - | - | - | 465 | 10496 | 522 | 7141 |
| Ischaemic stroke | - | - | - | - | 155 | 10870 | 171 | 7600 |
| Mortality | - | - | - | - | 428 | 10974 | 414 | 7702 |

The epigenetic index of pubertal development (epigenetic puberty) was correlated with chronological age in both cohorts and both sexes. In the HRS the correlation was moderate and similar across sexes (r=0.51, 95% CI 0.48 to 0.54 in females; r=0.51, 95% CI 0.47 to 0.55 in males), and weaker than for PCGrimAge (r=0.90 and r=0.88) or PhenoAge (r=0.73 in both); DunedinPACE, which captures the pace rather than the level of aging, correlated only weakly with age (r=0.08 and r=0.13). In GS, the correlation was substantially stronger (r=0.82 in females; r=0.86 in males), but among GS participants aged 55 and older it returned to the level seen in the HRS (r=0.50 and r=0.44), indicating that the difference reflects age span rather than a difference between cohorts. All subsequent analyses residualized epigenetic puberty, PhenoAge, and PCGrimAge on chronological age to index age acceleration, and all models were additionally adjusted for chronological age to account for potential cohort effects.

We then tested whether age-accelerated epigenetic puberty predicted aging-related biomarkers and health outcomes based on the mortality process model (Zhao & Crimmins, 2022), in sex-stratified models adjusted for chronological age (**Figure 2**; **Supplementary Table 1**). HRS outcomes included the epigenetic clocks, clinical-biomarker-based biological age (ExpBioAge, a biological age measure based on 22 clinical biomarkers, including physiological dysregulation), multimorbidity, functional limitations (ADL/IADL), cognitive dysfunction and decline, BMI, age at menopause, and 6-year mortality; GS outcomes included the epigenetic clocks and the 10-year onset of five aging-related disease groups and all-cause mortality, ascertained from linked national health records. In both cohorts and both sexes, more advanced epigenetic puberty acceleration was associated with accelerated epigenetic clocks (HRS b=0.13 to 0.47; GS b=0.05 to 0.40; all P<0.001, except PCGrimAge acceleration in HRS males, b=0.08, P=0.051), with clinical-biomarker-based biological aging in the HRS (ExpBioAge acceleration b=0.16 in females and b=0.15 in males), and with mortality (HRS 6-year mortality OR=1.46 and OR=1.39; all P<0.005). Additionally, among females, epigenetic puberty acceleration was significantly associated with higher multimorbidity, more ADL/IADL limitations, greater cognitive dysfunction and higher BMI (brange=0.06 - 0.16; all P<0.01) in HRS, and higher risk of 10-year incident diabetes, any cancer, lung disease and mortality (HRrange=1.12 - 1.19; all P<0.005) in GS. Among GS males, only lung disease reached significance (HR=1.18, P=0.034).

**Figure 2.**
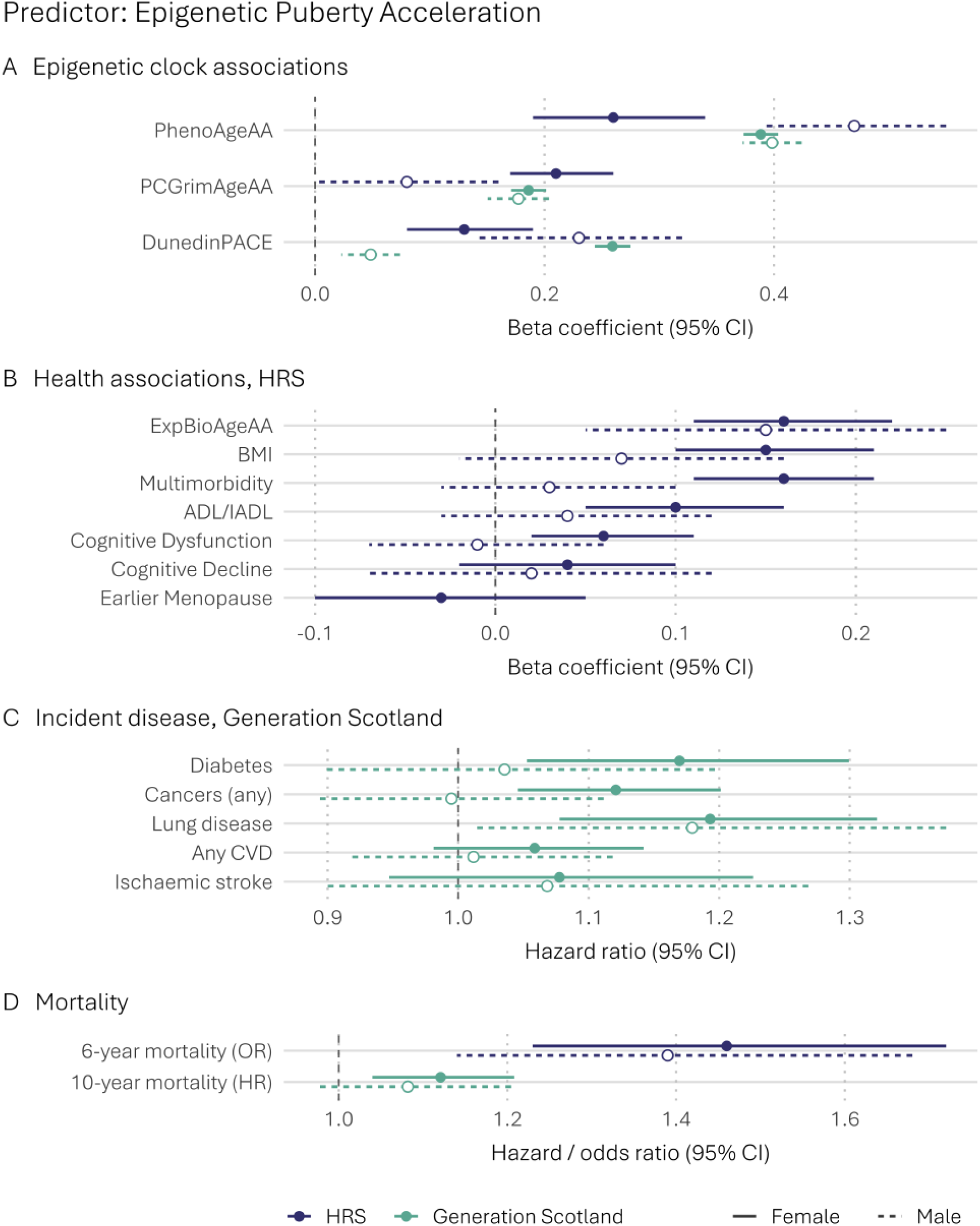
Associations between Age-Accelerated Epigenetic Puberty (DNAmPubertalAge Acceleration) and Aging Outcomes in the HRS and GS. All models are stratified by sex and adjusted for chronological age. HRS: Health and Retirement Study. GS: Generation Scotland. PhenoAge, GrimAge, and DunedinPACE are DNA-methylation-based biological aging measures. PC: Principal Component Version. ExpBioAge: Expanded Biological Age. ExpBioAge is based on clinical biomarkers to measure physiological dysregulation. AA: Age Acceleration. The AA version of an index was calculated using the residual that resulted from regressing the index on chronological age. ADL+IADL: Limitations of Activities of Daily Living and Instrumental Activities of Daily Living. CVD: Cardiovascular Diseases.

### 2. Epigenetic puberty index explains variation in aging outcomes in addition to epigenetic clocks

We next asked whether epigenetic puberty captured variation beyond the clocks, regressing each outcome on age-accelerated epigenetic puberty, one epigenetic clock at a time, and chronological age (**Supplementary Table 2**). Among HRS females, every association remained statistically significant after adjustment for all three clocks (all P<0.01), with the standardized coefficients for continuous outcomes reduced by 14% to 49% and the mortality odds ratio by 4% to 13%. Among HRS males, the association survived adjustment for PCGrimAge acceleration only for ExpBioAge acceleration (b=0.14, P=0.003) and 6-year mortality (OR=1.33, P=0.006). In GS, it persisted only for incident any cancer among females, and only with adjustment for PCGrimAge acceleration (HR=1.08, P=0.026) and DunedinPACE (HR=1.07, P=0.047).

For those models that survived clock adjustment, we decomposed the model R-squared using the Shorrocks-Shapley approach (**Figure 3**; **Supplementary Table 3**). Epigenetic puberty consistently explained less variation than the clock it was compared against, and considerably more in the HRS than in GS. Among HRS females it uniquely accounted for 2.7% to 3.4% of the variance in ExpBioAge acceleration, 2.1% to 2.4% in multimorbidity, 0.9% to 1.2% in BMI, 1.5% to 1.8% in 6-year mortality and 0.5% to 0.6% in ADL/IADL limitations; among HRS males, 0.5% to 0.8% for ExpBioAge acceleration and 0.2% to 0.4% for mortality. As a share of the total variance explained, this ranged from 5.8% to 40.4% in the HRS, but was only 1.2% to 1.3% for incident cancer in GS, where chronological age dominated the models (**Supplementary Figure 1**).

**Figure 3.**
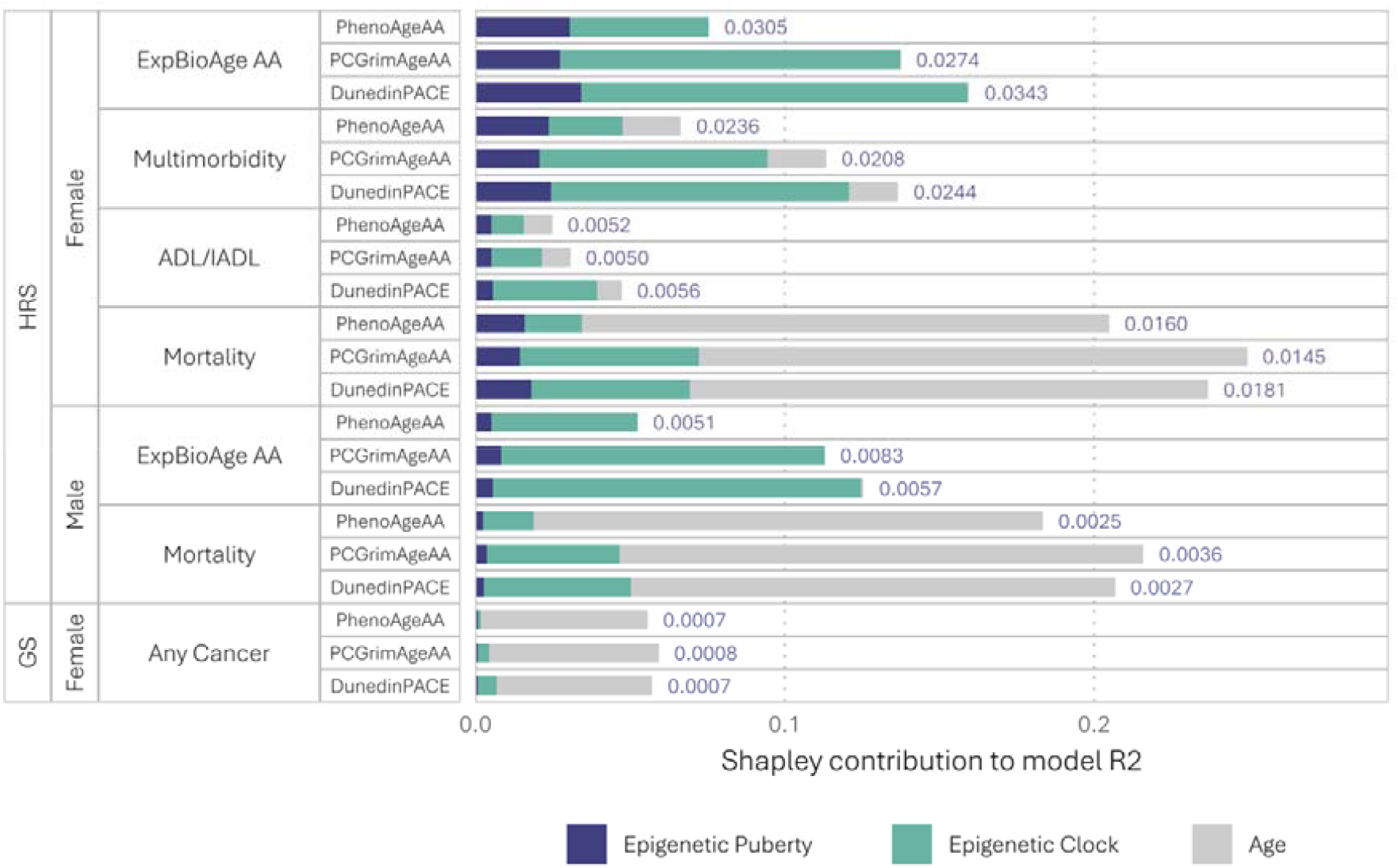
Unique Variance in Aging Outcomes Accounted for by Age-Accelerated Epigenetic Puberty (Blue), Epigenetic Clocks (Green), and Chronological Age (Grey), in the HRS and GS. HRS: Health and Retirement Study. GS: Generation Scotland. PhenoAge, GrimAge, and DunedinPACE are DNA-methylation-based biological aging measures. PC: Principal Component version. ExpBioAge: Expanded Biological Age, based on clinical biomarkers to measure physiological dysregulation. AA: Age Acceleration; the AA version of an index was calculated using the residual that resulted from regressing the index on chronological age. ADL+IADL: Limitations of Activities of Daily Living and Instrumental Activities of Daily Living. Each outcome was regressed on DNAmPubertalAge acceleration, one epigenetic clock at a time, and chronological age. The Shorrocks-Shapley approach was then used to decompose the model R-squared into the unique variance explained by each of the three variables. Outcomes shown are those for which the DNAmPubertalAge acceleration effect remained statistically significant after adding a clock; where this held for only one clock, all three clocks are shown. R-squared is the OLS R-squared for continuous outcomes and McFadden’s pseudo R-squared for the mortality logistic regression model in HRS, and Nagelkerke’s pseudo R-squared for the Cox models in GS.

### 3. Adulthood recalled earlier menarche and the epigenetic puberty index are not associated with each other and they capture aging outcomes differently

GS also collected adult recall of age at menarche, allowing the epigenetic puberty index to be compared with a phenotypic marker of pubertal timing in the same females. Age-accelerated epigenetic puberty was not associated with recalled age at menarche (b=0.01, P=0.26). Because the epigenetic index could be measured decades after menarche for some participants, we tested whether the recall interval mattered: the interaction with age at DNAm collection was not significant (P=0.30), and remained null after allowing for a quadratic effect of age (P=0.82).

To compare epigenetic and phenotypic puberty, recalled menarche was reverse-coded and standardized so that higher values indicate earlier menarche (**Figure 4**; **Supplementary Table 4**). Both predicted accelerated epigenetic aging on all three clocks, but associations were weaker for recalled menarche (b=0.02 to 0.05) than for epigenetic puberty (b=0.19 to 0.39); For incident disease, earlier menarche predicted diabetes (HR=1.30), any CVD (HR=1.11) and mortality (HR=1.11; all P<0.03) but not cancer, lung disease or stroke, whereas epigenetic puberty predicted diabetes, cancer, lung disease and mortality but not CVD.

**Figure 4.**
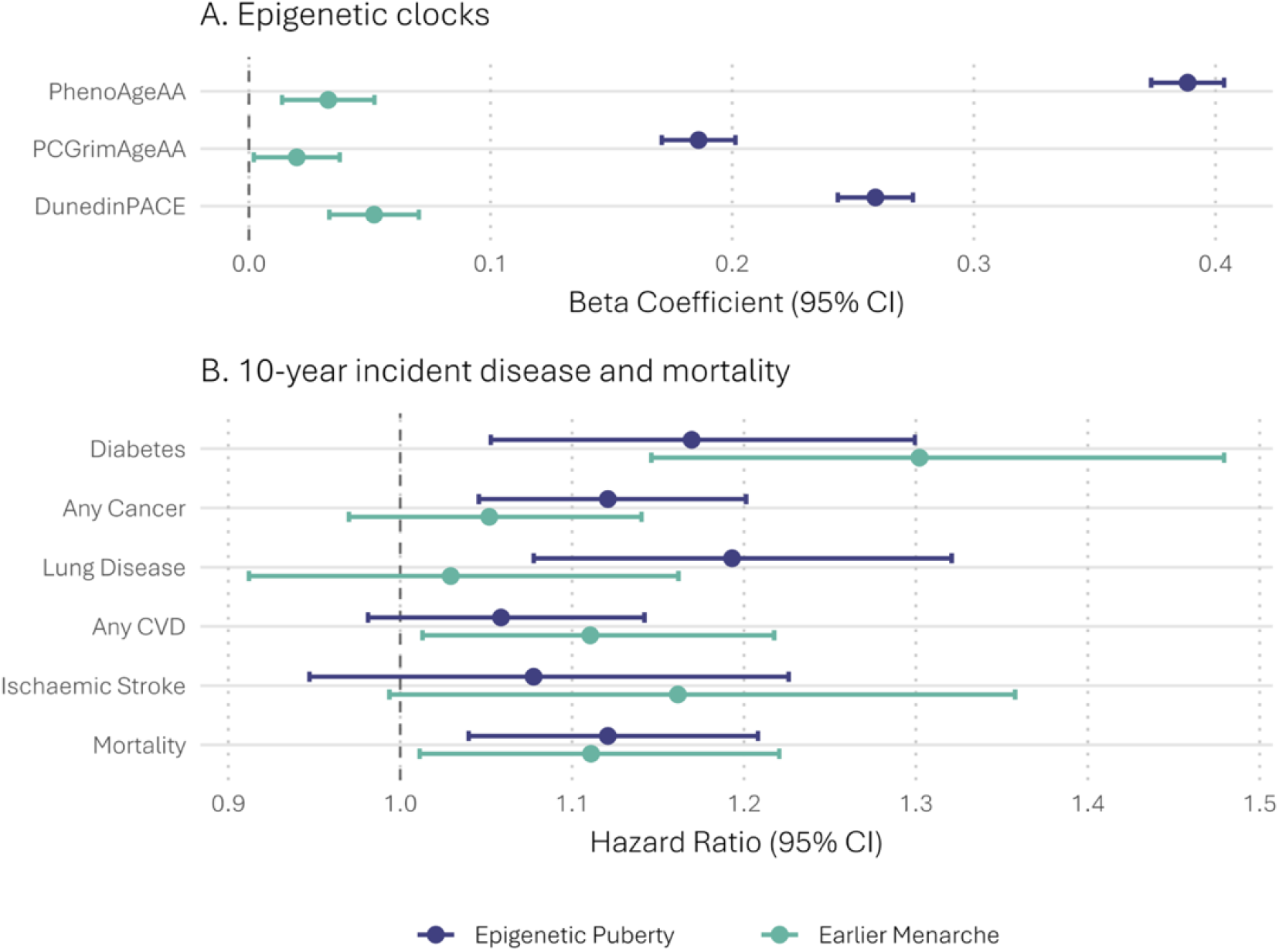
Associations of the Age-Accelerated Epigenetic Puberty Index and Adulthood-Recalled Earlier Menarche with Epigenetic Clocks and 10-Year Incident Disease among Females in GS. All models are adjusted for chronological age. Both epigenetic puberty and earlier menarche are standardized for comparison. PhenoAge, GrimAge, and DunedinPACE are DNA-methylation-based biological aging measures. PC: Principal Component version. AA: Age Acceleration; the AA version of an index was calculated using the residual that resulted from regressing the index on chronological age. CVD: Cardiovascular Diseases.

### 4. Adjusting for immune cell composition attenuates the observed associations

Because the epigenetic indices were derived from whole blood, we added the proportions of six major immune cell types to the models predicting phenotypic aging outcomes (**Supplementary Table 5**). In the HRS, associations of epigenetic puberty were substantially attenuated: ExpBioAge acceleration fell from b=0.16 to b=0.05 among females (P=0.077) and from b=0.15 to b=0.07 among males (P=0.168), and 6-year mortality from OR=1.46 to OR=1.22, while multimorbidity (b=0.11), ADL/IADL limitations (b=0.07) and BMI (b=0.14) remained significant but weaker; clock coefficients were attenuated as well but to a lesser degree. By contrast, in GS, associations with incident disease among females were unattenuated or slightly stronger (diabetes HR=1.25, any cancer HR=1.15, lung disease HR=1.27; all P<0.002), and only mortality (HR=1.12 to HR=1.05, P=0.240) and lung disease among males (HR=1.18 to HR=1.05, P=0.577) were reduced to the null. Immune cell composition therefore accounts for a substantial part of the link between epigenetic puberty and general aging phenotypes in the HRS, but little of its link with incidence of specific diseases in GS, consistent with the view that shifts in immune cell distribution are themselves part of biological aging (e.g., López-Otín et al., 2023).

## Discussion

In this study we found convergent evidence that the molecular pattern previously found to index accelerated pubertal development in adolescents is related to adult biological aging and health. First, individuals who were chronologically older in both the HRS and GS had DNA methylation patterns that more closely resembled those of adolescents who were more advanced in secondary sex characteristic development, as indicated by epigenetic puberty. The strength of this association was comparable to that observed for second-generation epigenetic clocks (i.e., PhenoAge and PCGrimAge). These findings provide initial evidence that the epigenetic index of pubertal development and indices trained on multisystem biomarkers of biological aging to some degree capture shared epigenetic processes across the lifespan (Ellis & Giudice, 2019; Gladyshev, 2021; Horvath & Raj, 2018; Raffington, 2024).

Second, we found that across both sexes and cohorts, advanced epigenetic puberty (residualized for chronological age) was associated with accelerated aging and increased mortality risk. In females from HRS, advanced epigenetic puberty was further associated with multimorbidity, functional limitations, cognitive dysfunction, and BMI, though not with cognitive decline. We extended these findings by utilizing the GS cohort to assess prospective disease onset over 10 years, going beyond the cross-sectional multimorbidity measure available in the HRS. Prospective disease incidence has become an important criterion for evaluating epigenetic aging biomarkers, because it separates the prediction of future health from the concurrent burden of existing illness (Mavrommatis et al., 2025). On this criterion, the epigenetic puberty index behaves like an aging biomarker not only among older adults but across adulthood more broadly. These findings mirror previous evidence that epigenetic puberty correlates with aging clocks and involves genes linked to longevity and aging, such as *FOXO3, BNF*, and *LHFPL* (deSteiguer et al., 2025), and are consistent with literature linking early pubertal timing to later-life disease and mortality (Elks et al., 2013; Lakshman et al., 2009; Ohlsson et al., 2020).

Third, epigenetic puberty accounted for a unique, albeit smaller, proportion of variance in downstream aging-related outcomes beyond that explained by epigenetic clocks. This was most evident in the HRS, where the association survived adjustment for every clock among females and, for biomarker-based biological age and mortality, among males; in GS it persisted only for incident cancer. This contrast likely reflects what each outcome captures: the GS outcomes are incident diagnoses of specific pathologies, whereas the HRS outcomes index more general or aggregated aging outcomes. This suggests that the underlying biology of pubertal development may add modest but distinct information for understanding and quantifying biological aging in general rather than disease-specific risk. Although cellular aging is thought to be a lifelong process that may commence very early in life (Gladyshev, 2021; Horvath & Raj, 2018; Raffington, 2024), few algorithms have considered processes in children and adolescents as relevant to the development of aging algorithms. The biological consequences of accelerated maturation could include heightened physiological stress and a reduced capacity for long-term maintenance, traceable in the epigenome (Belsky & Shalev, 2016; Ellis & Giudice, 2019). Consequently, generating and tracking biomarkers of development and biological aging from infancy onward represents a promising avenue for life-course epidemiology (e.g., PROSPER Consortium, Strauss et al., 2026).

Fourth, the magnitude of the associations between epigenetic puberty and aging were substantially reduced after adjusting for immune cell distribution in the HRS, whereas in GS the associations with incident disease were largely unchanged. Because the immune system undergoes significant functional maturation and structural reorganization during pubertal development (Resztak et al., 2023), and given that the subsequent age-related deterioration of immune function (immunosenescence) is an intrinsic process of biological aging (López-Otín et al., 2023), this immune cell subtype distribution may lie on the causal pathway between molecular-level aging biomarkers and health. That the attenuation was limited to the HRS could be explained by the same distinction mentioned above: immunosenescence is intrinsic to general biological aging, whereas the contribution of immune cell distribution to the onset of any specific disease is likely more complex and pathology-specific. This attenuation supports the interpretation that the biology of pubertal development may partially exert its influence on health through inflammatory and immune pathways (Ellis & Giudice, 2019).

The current study has limitations. First, lacking longitudinal epigenetic and pubertal measures, we cannot determine whether the adult methylation patterns of pubertal maturation accurately reflect developmental history or remain stable when measured repeatedly from adolescence onward. The GS data allowed a partial test of this: both epigenetic puberty and adulthood-recalled earlier menarche predicted the onset of aging-related disease, consistent with the existing literature (Elks et al., 2013; Lakshman et al., 2009). However, the two were not associated with each other among females and they predicted different disease outcomes. We therefore have no evidence that the epigenetic index accurately reflects past pubertal events. However, the objective of computing this index among adults was not to retrospectively infer pubertal timing. Instead, it was to detect and quantify the underlying biology of advanced adolescent puberty during the life stage when aging outcomes are more prevalent. In this sense, our study offers a transferable methodological framework: deriving a portable epigenetic marker that captures time-sensitive signatures of a developmental process, and probing that signature in later-life cohorts to understand its long-term health consequences (Raffington, 2024).

A further limitation is that the sample used to train the epigenetic puberty index was smaller than is typical for adult DNAm studies, even though the index used here were built from the largest DNAm study of pubertal development to date (deSteiguer et al., 2025). Consequently, the index may not fully capture the epigenetic signature of pubertal development, and observed associations likely represent a lower bound of detectable effects. Larger pediatric DNAm studies with more comprehensive pubertal phenotyping are needed to develop biomarkers that more accurately reflect underlying epigenetic signals.

In conclusion, these findings indicate that an epigenetic index of pubertal development behaves similarly to a biomarker of aging in adulthood by tracking chronological age, correlating with other measures of biological aging, and predicting aging-related morbidity and mortality. Because pubertal maturation is energetically costly and generates oxidative and inflammatory stress through rapid biosynthetic and endocrine activation, this accelerated developmental biology may function directly as a pro-aging mechanism. Notably, among females, this index provides incremental explanatory value beyond existing epigenetic clocks. These findings demonstrate that the measurement of cellular aging may benefit from considering developmental processes in early ontogeny.

## Methods

### Data and Participants

*The HRS (Health and Retirement Study) –* The HRS is a nationally representative longitudinal survey of individuals over age 50 in the United States. Blood DNAm was measured on a subsample of the 2016 Venous Blood Study component. A total of n=4,018 DNAm samples passed the quality control process (QC). The preprocessing of DNAm data in HRS can be seen in the **Supplementary Methods**. Survey weights computed by the HRS team for the VBS innovative sub-sample are used to adjust for initial sample selection and differential consent and completion. After applying weights, the sample is representative of community-dwelling older adults in the US.

Our main analytical sample consisted of 3,642 respondents (2,106 females and 1,536 males), who had a valid VBS innovative sub-sample survey weight, and who further had complete information on sociodemographic characteristics, multimorbidity, ADL & IADL functional limitations, and cognitive function. The models for some specific aging outcomes were based on non-missing subsamples (**Table 1)**.

*The GS (Generation Scotland) cohort –* GS is a population- and family-based research study comprised of 24,096 volunteers. Volunteers joined the cohort between 2006 and 2011 with the majority of recruitment taking place via invitation from the individuals’ general practitioners, and the majority agreed to provide blood, from which epigenetic data was profiled (Walker et al., 2025). A total of n=18,869 DNAm samples passed the QC process. Our analytical sample for the GS secondary analysis consisted of 18,676 participants (10,974 females and 7,702 males), who had valid DNAm data and information on 10-year onset of a selected set of aging-related diseases. The preprocessing of DNAm data in GS is described in the Supplementary Methods, and the demographic characteristics, 10-year incident cases, and number at risk of selected diseases can be seen in **Table 1**.

### Measures

#### Epigenetic index of pubertal development

The epigenetic index of pubertal development, DNAmPubertalAge, is an elastic-net-based index of pubertal development, trained to predict pubertal age, which is the age in years at which the child’s reported pubertal development is the norm. (For example, if an 11-year-old girl reported secondary sex characteristic development that was typical of 14-year-olds, then her pubertal age would be 14 years). The index had high measurement reliability: The intraclass correlation coefficient was 0.88 (95% CI 0.55 to 0.97) among 8 technical replicates in girls and 0.87 (95% CI 0.59 to 0.96) among 11 technical replicates in boys (deSteiguer et al., 2025). To measure pubertal development beyond the effect of chronological age, the age-accelerated version of the epigenetic puberty index (DNAmPubertalAge acceleration) was calculated using the residual that resulted from regressing DNAmPubertalAge on chronological age. Although the epigenetic puberty index was generated in adolescents’ saliva samples, it yields acceptably moderate saliva-blood correspondence (intraclass correlation coefficient, 0.64) (**Supplementary Methods and Supplementary Table 6**). Data from the 2016 venous blood collection of HRS were used for epigenetic index computation.

#### Multiple levels of aging outcomes in HRS

To leverage the extensive health information in HRS and measure a full spectrum of aging outcomes, multiple sequential and interrelated dimensions of aging (Zhao & Crimmins, 2022) were used in the current study, including (1) molecular-level aging (measured by epigenetic clocks), (2) physiological dysregulation (measured by physiological biological age measure based on clinical biomarkers), (3) diseases and conditions (measured by multimorbidity), (4) functioning loss (measured by ADL and IADL functional limitations, cognitive dysfunction, and cognitive decline), and (5) mortality (6-year mortality). Details for these outcomes are included in **Supplementary Methods**.

#### 10-year onset of aging-related diseases in GS

GS participants consented to the linkage of their study data to national electronic health records, from which disease outcomes were derived using consensus definitions based on primary and secondary care codes. Although participants consented to the linkage of their primary care records, these records were available for only about 40% of the cohort, because access additionally required agreement from each general practice as the data holder (Mavrommatis et al., 2025). Mortality was ascertained through Community Health Index linkage to the National Records of Scotland. To parallel the multimorbidity measure used in HRS, five groups of aging-related diseases were selected, including (1) diabetes, (2) cancers (any), (3) lung disease, (4) any cardiovascular disease, and (5) ischaemic stroke, as well as all-cause mortality. Participants with a diagnosis recorded before the blood draw were excluded from the analysis of that disease, so that all outcomes reflect incident cases. Time to onset was measured from the blood draw over 10 years of follow-up. 10-year incident cases and number at risk of selected diseases can be seen in **Table 1**.

#### Major leukocyte subtypes

The HRS immune cell type proportions were evaluated using flow cytometry performed on peripheral blood mononuclear cells (PBMCs) isolated from whole blood. In the models with cell type adjustment, the percentages of granulocytes, natural killer cells, B cells, CD4+ T cells, CD8+ T cells, and monocytes were adjusted (Salas et al., 2018). In GS, proportions of the same 6 cell types were estimated by DNA methylation data using the EPIDISH package (Teschendorff et al., 2017).

### Statistical Analysis

The preregistration of the analyses can be found at https://doi.org/10.17605/OSF.IO/FXSV7. Since preregistration, we have additionally acquired access to the GS cohort, so unpreregistered GS analyses were further added. Survey weights for the HRS VBS innovative sub-sample were used for all HRS estimates and models to adjust for initial sample selection and differential consent and completion. All models were stratified by sex and adjusted for chronological age; no further covariates were included. The epigenetic puberty index, the epigenetic clocks and all continuous outcomes were standardized. Analyses were performed using Stata version 18 and R version 4.4.

First, we examined associations between age-accelerated epigenetic puberty and chronological age, and between age-accelerated epigenetic puberty and each aging outcome, separately for each sex and cohort. Continuous outcomes were modelled with linear regression, 6-year mortality in the HRS with logistic regression, and the 10-year onset of aging-related diseases and mortality in GS with Cox proportional hazards models.

Second, we examined whether age-accelerated epigenetic puberty accounted for unique variation in aging beyond the epigenetic clocks. Each clock was added, one at a time, to the model relating epigenetic puberty to the outcome, so that every model contained three variables: epigenetic puberty, one clock, and chronological age. For those outcomes in which the epigenetic puberty association remained significant after adding at least one clock, the Shorrocks-Shapley approach was then used to decompose the model R-squared into the variance uniquely attributable to each of the three variables; where the criterion was met for only one clock, all three clocks are reported for comparability. R-squared is the ordinary least squares R-squared for continuous outcomes and McFadden’s pseudo R-squared for mortality in the HRS, and Nagelkerke’s pseudo R-squared for the Cox models in GS; the HRS decompositions are unweighted, as the decomposition does not accommodate survey weights.

Third, we compared the epigenetic puberty index with a phenotypic marker of pubertal timing available in GS, adult recall of age at menarche. We tested whether age-accelerated epigenetic puberty predicted recalled menarche, whether that association was moderated by age at DNA methylation collection, and how the two markers compared in their associations with the epigenetic clocks and in predicting incident disease. Recalled menarche was reverse-coded and standardized so that higher values indicate earlier menarche, matching the direction and scaling of age-accelerated epigenetic puberty.

Fourth, as a sensitivity analysis, the proportions of six major leukocyte subtypes were added as covariates to the models in which an epigenetic index predicted a phenotypic aging outcome, to test whether the observed associations were attributable to between-person differences in immune cell composition.

## Data Availability Statement

Much of the Health and Retirement Study (HRS) data can be accessed via the University of Michigan site using the online application system (https://hrs.isr.umich.edu/data-products). Sensitive health data from HRS is only available under restricted access: Venous Blood Study (VBS) and Biomarker Data used in the current study can be accessed using the HRS sensitive health data online application system (https://hrsdata.isr.umich.edu/data-products/sensitive-health). HRS Epigenetic data can be accessed through The National Institute on Aging Genetics of Alzheimer’s Diseases Data Storage Site (NIAGADS) upon application (https://hrs.isr.umich.edu/data-products/genetic-data). More information on the NIAGADS application can be found on its official website (https://niagads.scrollhelp.site/support/application-instructions).

Generation Scotland (GS) data are not publicly available, as they contain information that could compromise participant consent and confidentiality, and can be obtained from the data owners upon application. This includes the DNA methylation data and the linked electronic health record data used in the current study. Access requests can be made through the researcher portal (https://gsaccess.igc.ed.ac.uk), where they are reviewed and approved by the Generation Scotland Access Committee, and approved projects require a Data and Materials Transfer Agreement. Information on GS data and the access process can be found on the GS website (https://genscot.ed.ac.uk/for-researchers/). Applications from researchers in the UK and from international collaborators are both accepted. A searchable data dictionary describing the available phenotypic, clinical, and biochemical data can be found at https://datashare.ed.ac.uk/handle/10283/2988. Further questions for the GS team can be directed to the Generation Scotland Access Committee at.

## Funding

Max Planck Society (QW, YEW, SZ, LR)

National Institutes of Health grant R01HD114 d724 (LR, KPH)

European Union grant 101150809 (YEW)

Jacobs Foundation (LR, KPH)

European Union grant #101073237 (LR)

IMPRS Life (SZ, LR)

Biotechnology and Biological Sciences Research Council UKRI/BB/C001941/1 (SG, RM)

Generation Scotland (GS) was funded by a grant from the Chief Scientist Office of the Scottish Government Health Directorates (CZD/16/6) and the Scottish Funding Council (HR03006). Genotyping and DNA methylation profiling of the GS samples was carried out by the Genetics Core Laboratory at the Edinburgh Clinical Research Facility, University of Edinburgh, Scotland, and was funded by the Medical Research Council UK and the Wellcome Trust (Wellcome Trust Strategic Award ‘STratifying Resilience and Depression Longitudinally’ Reference 104036/Z/14/Z). DNA methylation profiling was also funded in part by the Wellcome Trust Investigator Award (Reference 220857/Z/20/Z). GS is currently supported by the Wellcome Trust (216767/Z/19/Z).

The Health and Retirement Study (HRS) is sponsored by the National Institute on Aging (grant number NIA U01AG009740) and is conducted by the University of Michigan.

## Author contributions

Conceptualization — QW, SG, AJD, SZ, YEW, TS, JAG, CM, EMC, KPH, RM, LR

Methodology — QW, SG, AJD, SZ, LR Formal analysis — QW, SG

Data curation — QW, SG, AJD, SZ Writing – original draft — QW, LR

Writing – review & editing — QW, SG, AJD, SZ, YEW, TS, JAG, CM, EMC, KPH, RM, LR

Visualization — QW, SG Supervision — LR

Funding acquisition — LR, RM, KPH

